# Discriminating cognitive performance using biomarkers extracted from linear and nonlinear analysis of EEG signals by machine learning

**DOI:** 10.1101/2020.06.30.20143610

**Authors:** K R Shivabalan, Deb Brototo, Goel Shivam, S Sivanesan

## Abstract

Nonlinear dynamics and chaos theory are being widely used nowadays in neuroscience to characterize complex systems within which the change of the output isn’t proportional to the change of the input. Nonlinear systems compared to linear systems, often appear chaotic, unpredictable, or counterintuitive, and yet their behaviour isn’t random. The importance of the time series analysis, which exhibits a typical complex dynamics, within the area of nonlinear analysis can’t be undermined. Hidden important dynamical properties of the physiological phenomenon can be detected by many features of these approaches. Nonlinear dynamics and chaos theory are being employed in neurophysiology with the aim to elucidate the complex brain activity from electroencephalographic (EEG) signals. The brain is a chaotic dynamical system and further, their generated EEG signals are generally chaotic in another sense, because, with respect to time, the amplitude changes continuously. A reliable and non-invasive measurement of memory load which will be made continuously while performing a cognitive task would be very helpful for assessing cognitive function, crucial for the prevention of decision-making errors, and also the development of adaptive user interfaces. Such a measurement could help to keep up the efficiency and productivity in task completion, work performance, and to avoid cognitive overload, especially in critical/high mental load workplaces like traffic control, military operations, and rescue commands. We have measured the linear and nonlinear dynamics of the EEG signals in subjects undergoing mental arithmetic task and measured the cognitive load on the brain continuously. We have also differentiated the subjects who can perform a mental task good and bad and developed a system using support vector machine to differentiate rest and task states.

---

Complex task performance requires the integration of knowledge related to the task, working memory, attention and decision making. Cognitive load refers to the amount of mental demand imposed by a particular task on these parameters^1^. A reliable, objective and cost effective method of assessing cognitive load is crucial in understanding the mental capacity of people. Experts have more knowledge or experience with regard to a specific task which reduces the cognitive load associated with the task in contrast to novices who have a heavier cognitive load^2^. Measuring and assessing the cognitive load associated with different tasks is crucial for many applications, monitoring the mental well-being of people who require heavy cognitive load in their occupation. Heavy cognitive load increases the chances of error in the task at hand^3–5^ or stereotyping^6^. Subjective measures of cognitive load^4^ may provide useful for cognitive load measurement in embodied scenarios if an appropriate survey is chosen. However, the use of cognitive load questionnaires has its own theoretical and practical issues. Different phrasings in cognitive load question items might lead to results that may not be comparable^7^. In addition to behavioral measures, cognitive workload has been assessed using physiological measures, such as pupil diameter^8^. The application of objectively determining the cognitive load has its growing application in ergonomics among the pilots or drivers.^9^

Electroencephalography is an easy, cost effective investigation to assess the brain activity. The focus of this paper is on quantifying cognitive workload using measures based on EEG. EEG-based measures for cognitive load have been described in several studies. The relationship among different spectral features to help predict cognitive load from EEG has been described in many studies^10–12^.

A wide range of methods have been applied for measuring and classifying the memory load using EEG signal. They can use either linear or nonlinear dynamic features to study them. Complex signals are information-rich. Another reason behind our preference to utilization of nonlinear measures in analysis of EEG is that nonlinear (nonstationary and irregular) signals nullify comprehensive understanding by a classic reductionist approach. Even the simplest nonlinear signals (originating from complex systems) will nullify the criteria of proportionality and superposition characteristic for linear systems^13^. The application of non-linear methods in classifying mental tasks is more recent, and measures like correlation dimension (CD)^14–16^, fractal dimension^17–19^, Hurst exponent (HE)^9,15,20^, sample entropy^18^, approximate entropy (ApEn)^15,21,22^ and largest Lyapunov exponent (LLE)^14,15^ have been used to measure the complexity/irregularity of the underlying brain dynamics during the performance of some cognitive tasks compared with the rest condition. Stated differently, in these studies the brain activity states; such as rest and stimulated have been differentiated. But to date, these measures have not been investigated in the analysis of the varying working memory load and the question whether these approaches could provide some information on discriminating subjects who can perform the arithmetic task well and subjects who can’t perform well.

In this study we have tried to understand the chaotic nature of the brain waves in the face of serial cognitive workload in normal subjects and discrimination of the ‘good’ and ‘bad’ performers when faced with increasing cognitive load.

In future, this study might be extrapolated to subjects with dementia or minimal cognitive impairment and will detect such degenerative processes at an earlier stage. It could also help in ensuring productivity in task completion and ergonomics.

## Results

### Compare rest with task state

First, we calculated the PSD, SE, FE and FD of five frequency bands (delta, theta, alpha, beta and gamma) and whole frequency signal. The alpha band changes from low-amplitude, non-rhythmic electrical activity to high-amplitude, oscillatory activity during the period of eye-closure^23^. We also calculated the ratio of features on alpha band to features on other bands (delta, theta, beta and gamma). The PSD results were shown in Figure 4.

**Figure 4.**
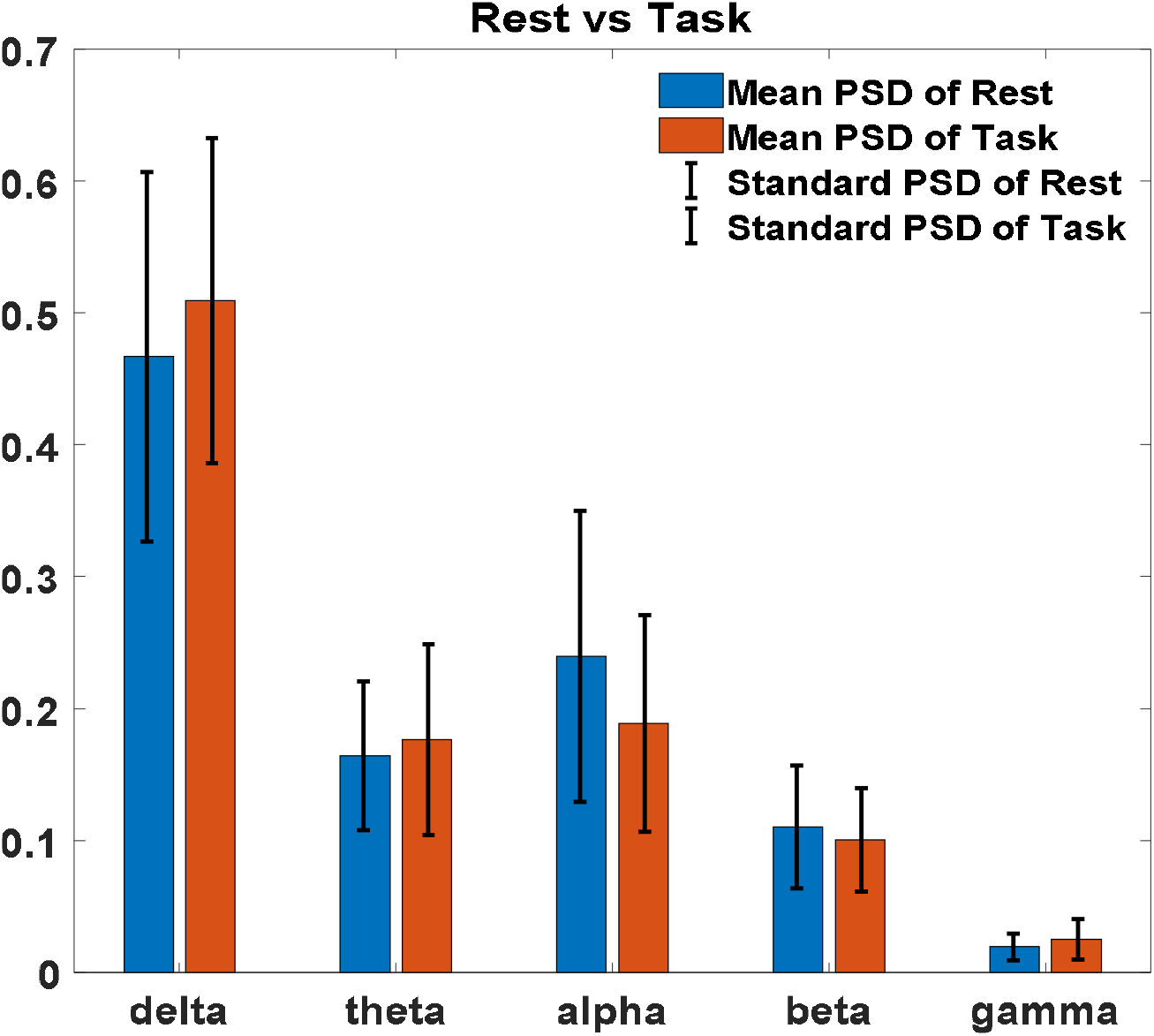
PSD of five frequency bands on FP 1 used to compare rest state with task state.

Then, the significant difference between rest and task states based on these features were also analyzed. First, we tested whether the data satisfied the assumption of normality and the assumption of homogeneity of variance. We chose the parametric test ANOVA1, which is a one-way analysis of variance for the data that satisfied the assumptions. Otherwise, a nonparametric test Kruskal-Wallis was used. It’s shown that the feature ***log(PSD of alpha/PSD of gamma)*** has the best performance and the results are shown in Figure 5. We found that the subjects had a higher alpha and lower gamma during rest state.

**Figure 5.**
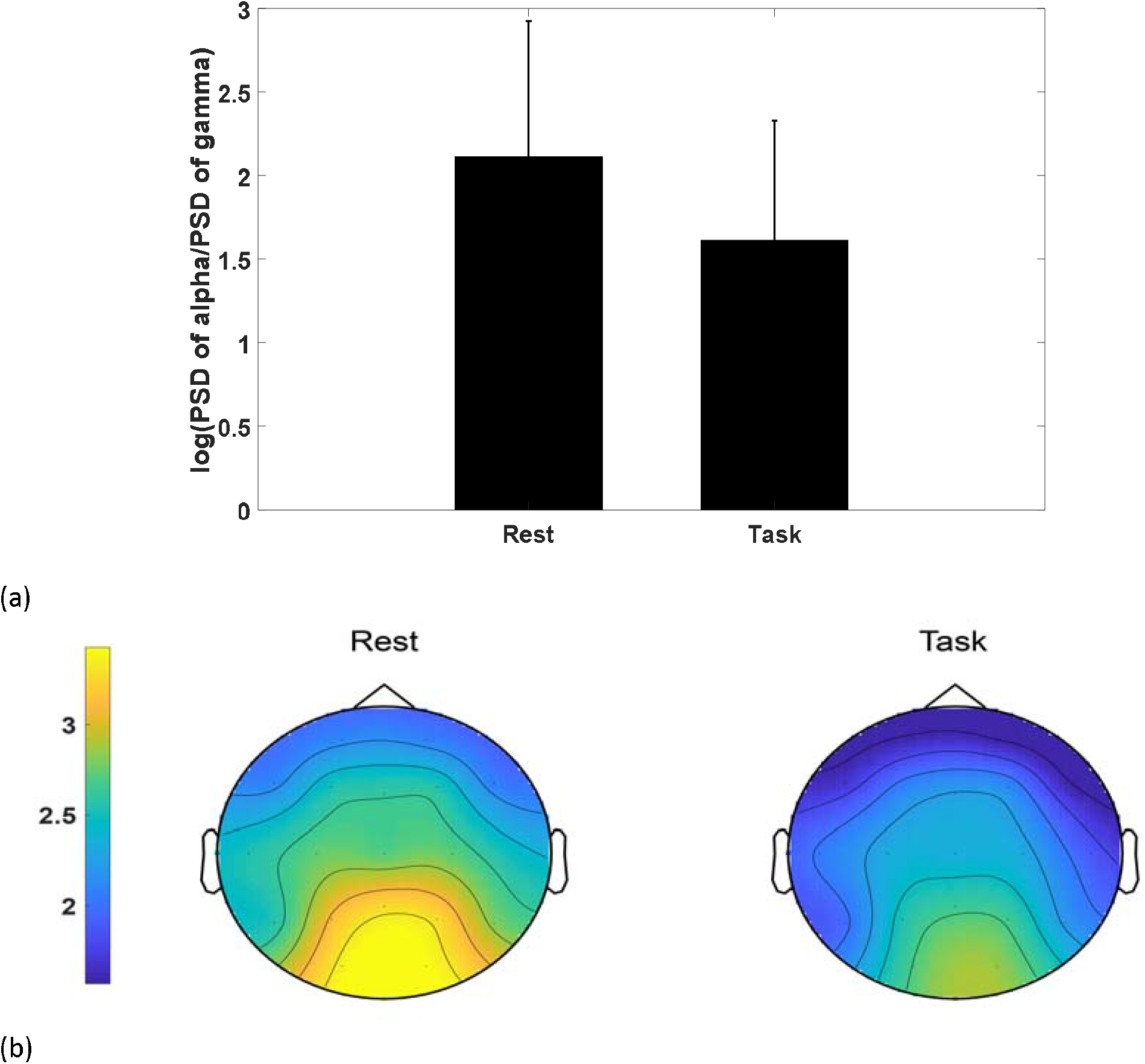
(a) the feature ***log (PSD of alpha/PSD of gamma)*** on FP1 for rest and task states (b) Spectral topographic map of power for ***log (PSD of alpha/PSD of gamma)*** for rest group and task group. Upper portion for each map shows the nasal and lower portion shows the occipital side. The color bar represents the log-transformed spectral power density (10*log10 (μv2 /Hz)) where yellow represents the maximum and blue represents the minimum values.

### Compare good with bad quality

First, we calculated the PSD, SE, FE and FD of five frequency bands (delta, theta, alpha, beta and gamma), ratio of alpha to each of the frequency band and ratio of the alpha to the whole frequency band. The PSD results were shown in Figure 6.

**Figure 6.**
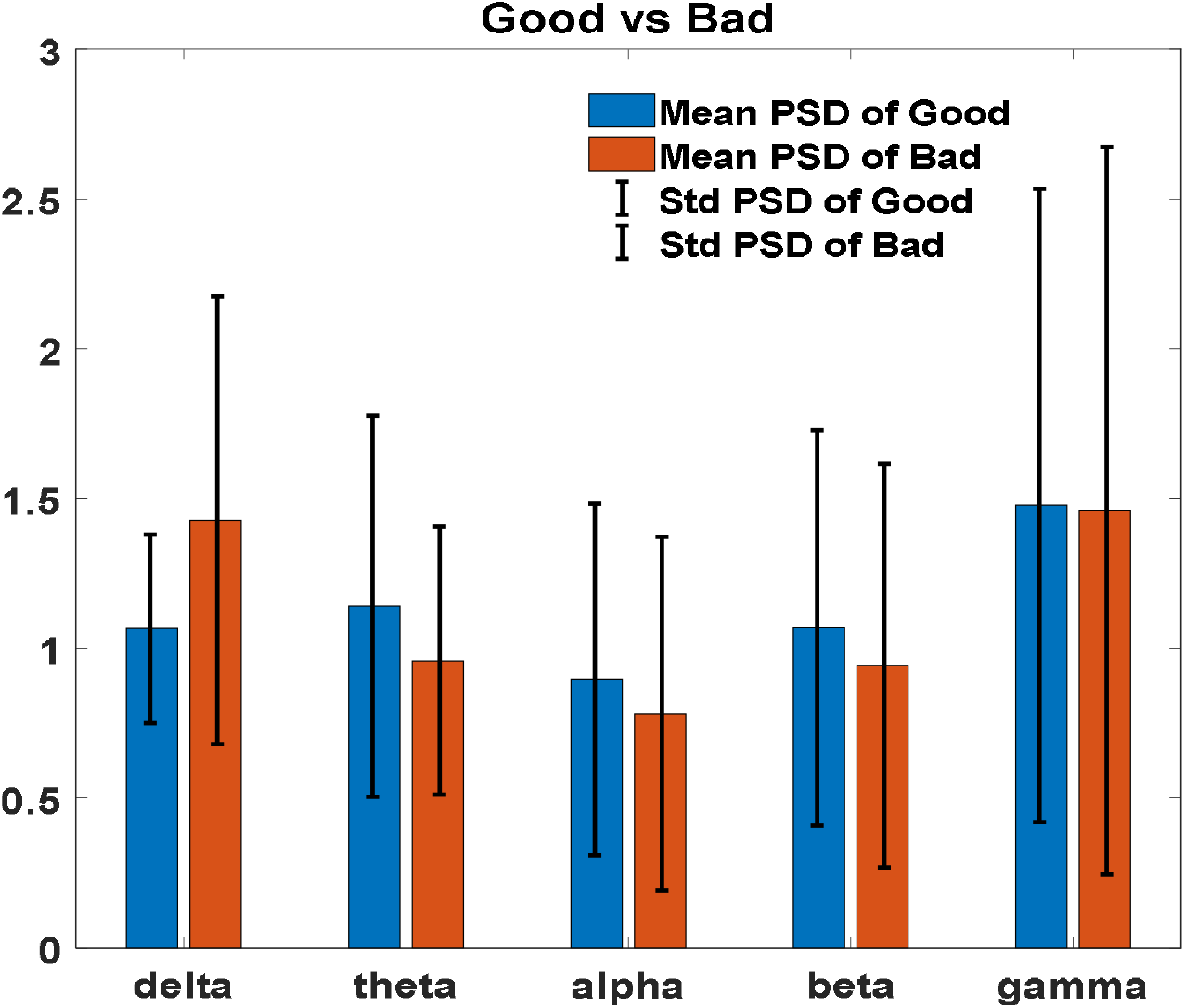
PSD of five frequency bands on FP 1 used to compare good quality with bad quality.

Theta and beta band oscillations directly reflect such cognitive processes. So we focused on theta and beta on PSD, SE, FE and FD, the results were shown in Figure 7 and Figure 8. As we can see that, the result is significant in frontal, temporal and occipital lobes. Good group has lower entropy and FD of theta and beta band in frontal lobe than bad group, while good group has higher entropy and FD of theta and beta in occipital lobe and temporal lobe. Good group has lower PSD of theta and beta bands in occipital lobe than bad group. Good group has higher PSD of theta and beta bands in left-frontal lobe than bad group.

**Figure 7.**
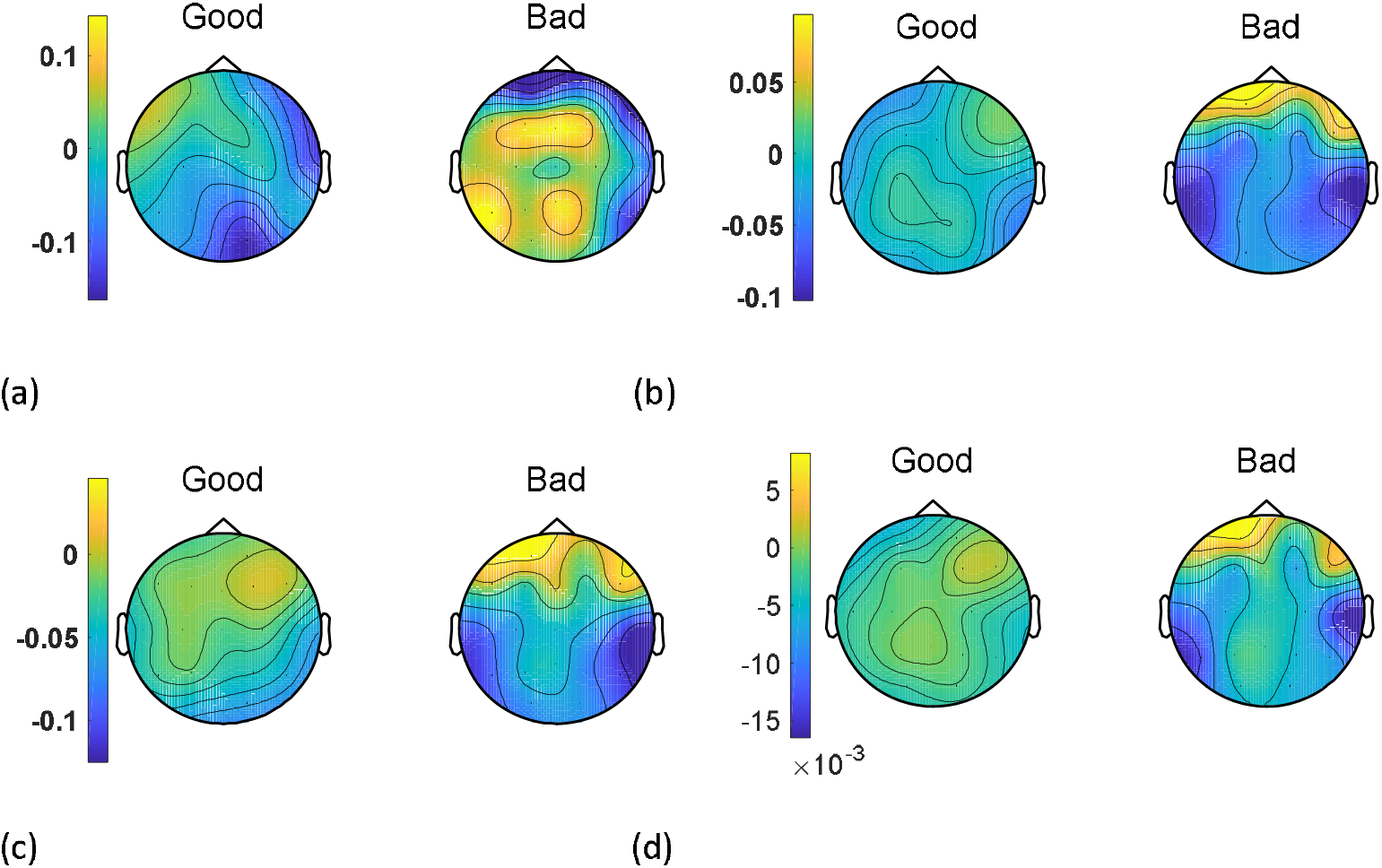
Spectral topographic map for theta band for good group and bad group. Upper portion for each map shows the nasal and lower portion shows the occipital side. The color bar represents the log-transformed spectral power density (10*log10 (µv2 /Hz)) where yellow represents the maximum and blue represents the minimum values. (a) PSD (b) SE (c) FE (d) FD.

**Figure 8.**
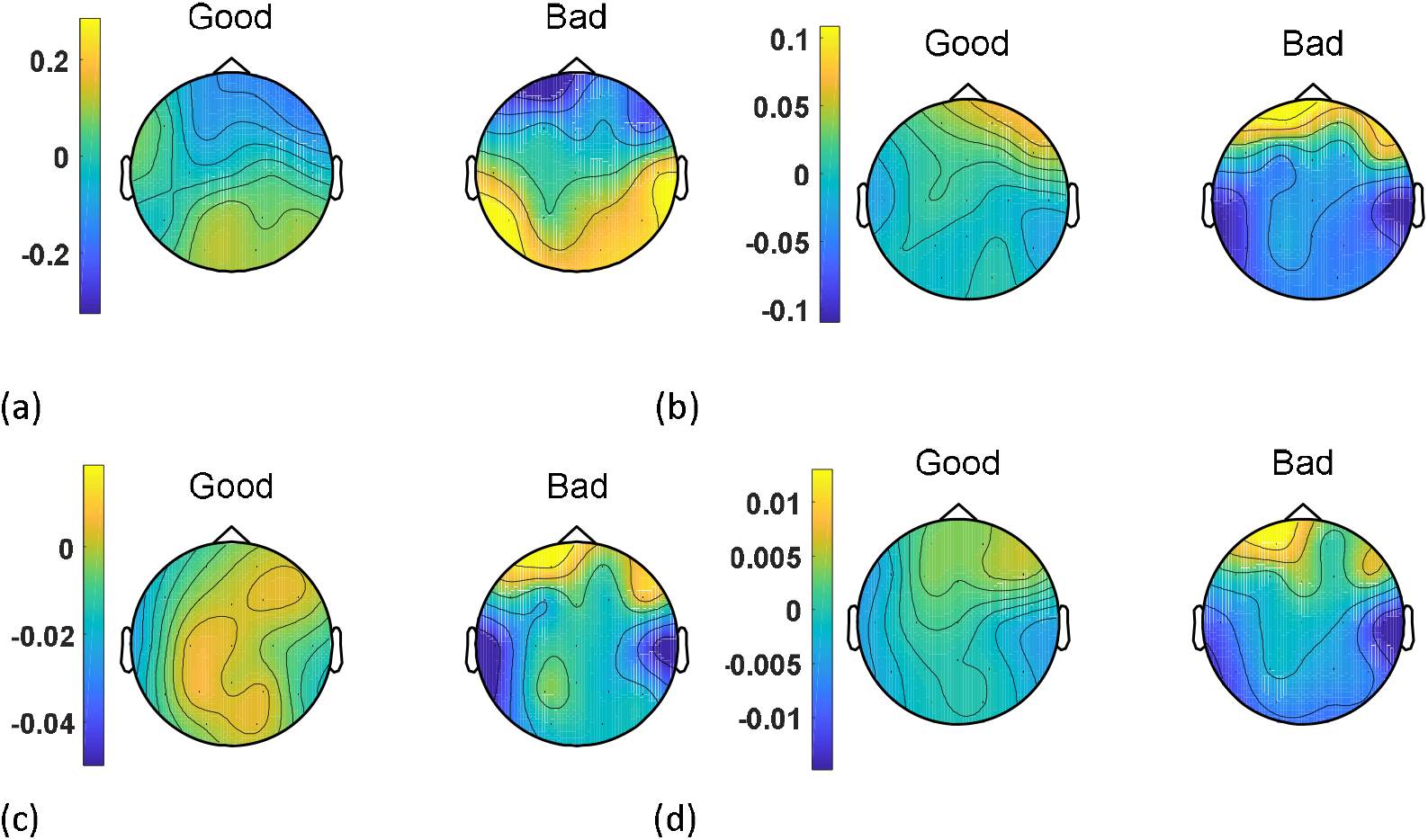
Spectral topographic map for beta band for good group and bad group. Upper portion for each map shows the nasal and lower portion shows the occipital side. The color bar represents the log-transformed spectral power density (10*log10 (µv2 /Hz)) where yellow represents the maximum and blue represents the minimum values. (a) PSD (b) SE (c) FE (d) FD.

### Classification

The extracted feature ***log(psd of alpha/psd of gamma)*** was fed into the Support vector machine (SVM), k-nearest neighbor (KNN) and Linear Discriminant Analysis (LDA) for classification of rest and task state in the MATLAB (R2018b) (MathWorks Inc., Natick, MA). The 10-fold cross-validation was used before LIBSVM to divide the samples into 10 parts. One of the 10 parts was used as a testing set and the remaining nine parts were used as a training set. To avoid information leakage, the samples from the same subject were divided into either training set or testing set. The k-fold cross validation method is used to divide the actual sample into k subsamples. Here k is chosen as 10. The mean value of 10-fold cross-validation will be used as the accuracy of this model. SVM produced the highest accuracy **85**.**31%**, while the accuracy of KNN and LDA were **83**.**44%** and **77.05%**.

Thereafter, we analyzed the performance of SVM in detail. The performance of the SVM for classification depends on the choice of a kernel. Optimization of parameters of a kernel can train our classifier for a given dataset and improve classification accuracy of a classifier. In this paper, the Polynomial Kernel is chosen. The degree of Polynomial Kernel has a direct influence on flexibility of resulting classifier^24^. The Polynomial Kernel is a global kernel which has a good generalization ability. Parameters of a kernel have a significant effect on the decision boundary. It can classify data with nonlinear boundaries as well as of high dimensions^24^. After the SVM provides the results, the confusion matrix, which contains **precision, recall, sensitivity**, and **specificity**, is constructed. The results were shown in Figure 9 and Table 3. The receiver operating characteristic curve (ROC) of SVM had an AUROC of 88.2% as shown in Figure 10.

**Figure 9.**
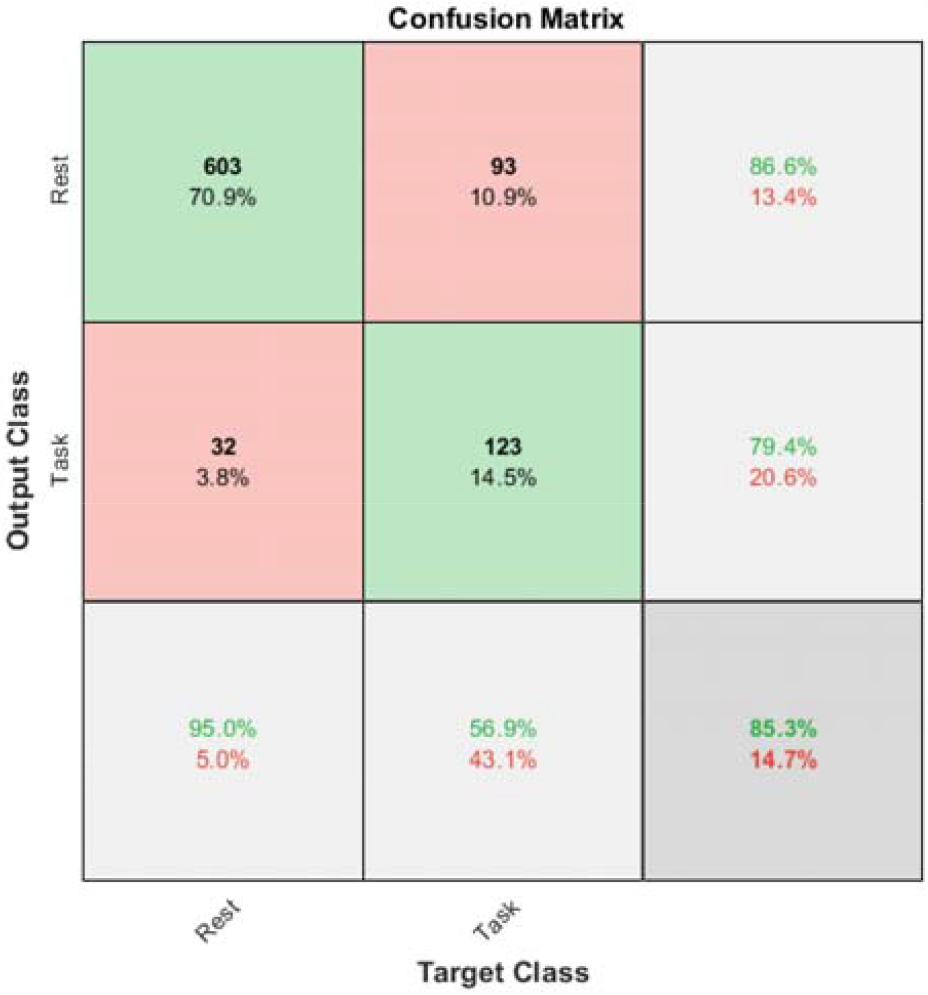
Support vector machine (SVM) polynomial kernel model confusion matrix

**Table 3.** Precision, recall/sensitivity, specificity and accuracy of SVM model

| Methods | Evaluations | Value |
| --- | --- | --- |
| Log(PSD of alpha/PSD of gamma) | Precision | 86.64% |
|  | Recall/Sensitivity | 94.96% |
|  | Specificity | 56.94% |
|  | Accuracy | 85.31% |

**Figure 10.**
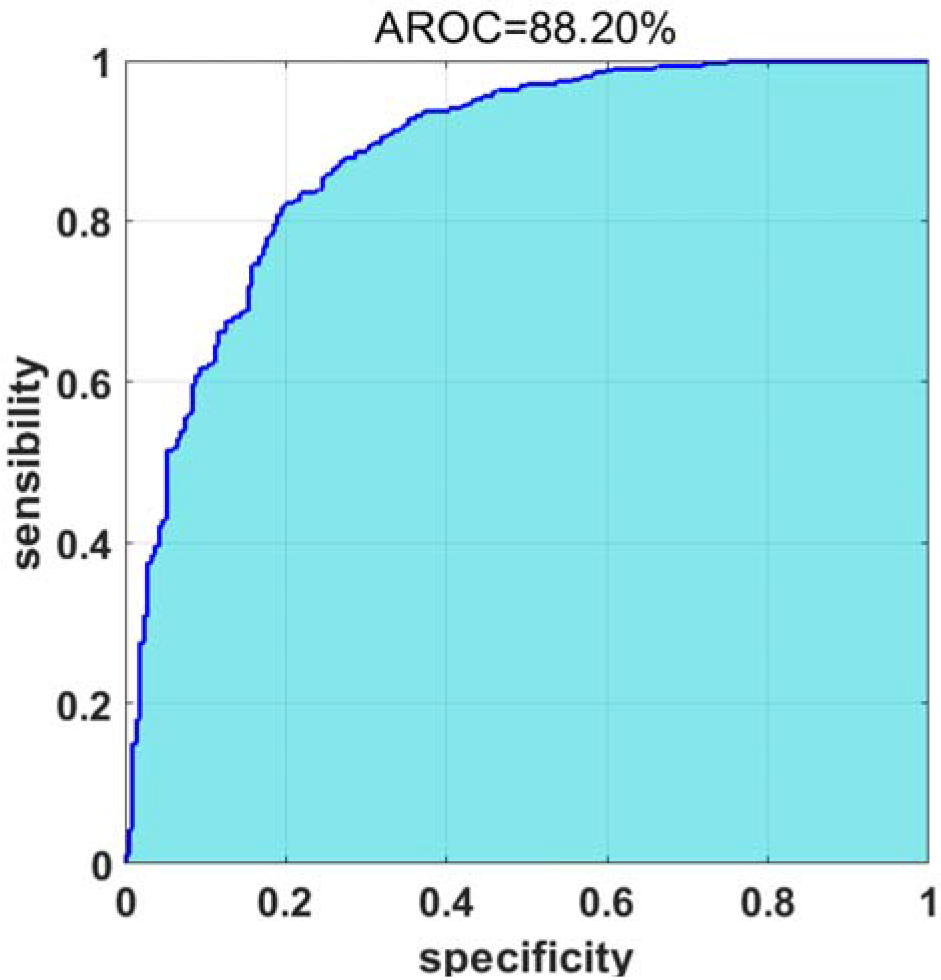
Receiver operating characteristic curve (ROC) of ***log(psd of alpha/psd of gamma)*** for discriminating rest and task state.

## Discussion

In this paper we have elucidated the chaotic nature of the brain waves in the face of serial cognitive workload in normal subjects and discrimination of the ‘good’ and ‘bad’ performers when faced with increasing cognitive load. The broader objective of this work is to develop a biomarker which can discriminate good performers from bad performers in a mental arithmetic task.

Beta and theta band oscillations directly reflected cognitive processes. This finding wasn’t very much congruent with the papers which reported that alpha and theta oscillations were reflective of cognitive and memory performance^25–27^. Although it was in congruence with the findings described by Molnar et al^28^. We observed that, there were significant differences among the frontal, temporal and occipital lobes. Good group had lower entropy of theta and beta band in frontal lobe but higher in occipital and temporal lobe when compared to the bad group. This implies that the complexity of the brain dynamics is less in the frontal and more in the occipital and temporal groups in complex cognitive processes. Or in other words, higher the mental work load on the memory, lower the entropy of the EEG signals in the frontal and higher in the occipital and temporal lobes. A decreased value of entropy with increased task load implies higher predictability and less irregularity in the brain activity. This finding is supported by Zarjam et al who showed that entropy decreases with increase in cognitive load mainly in the frontal lobe but it was more prominent in the delta sub band and they had measured approximate entropy and spectral entropy^20^. Both the studies show that brain behaves in a more focused and regular manner when performing more difficult tasks. Entropy correlated with fractal dimension positively. Or in other words Fractal dimension behaved similarly to that of entropy. Good group had lower entropy and FD of theta and beta band in frontal lobe than bad group, while good group had higher entropy and FD of theta and beta in occipital lobe and temporal lobe. While Wang et al showed that Fractal dimension increased with increased cognitive load mainly in the frontal lobe^29^. This contradiction couldn’t be explained by us. PSDs of theta and beta bands were higher in frontal but lower in occipital lobe when compared to bad group. This shows that higher PSD correlates with higher mental workload in frontal lobe. Zafar et al showed the involvement of delta oscillations in occipital region during cognitive tasks and they also showed that during the cognitive task the delta band has a different behavior compared to the rest task^30^. And they didn’t classify the task group into good and bad performers. This might be explained by the fact that their experiment was done with the subjects’ eyes’ closed.

But we have developed a marker which can differentiate rest and task states and the same was fed into SVM which had a greater predictive capacity. Due to the processed non-linear nature and a good generalizability of the feature set, the polynomial kernel function was used in our SVM model, as such approaches has been reported in similar situations. The feature set was subjected to various feature selection methods to narrow down more prominent features before using a SVM model. ***Log(psd of alpha/psd of gamma)*** had a better predictive capacity with a good area under ROC to discriminate rest from task states. SVM produced the highest accuracy **85.31%**, while the accuracy of KNN and LDA were **83**.**44%** and **77**.**05%**. Attallah used the same dataset to classify mental stress states and non-stress states using Principal Component analysis(PCA)^31^. PCA reduces the features extracted from such electrodes to lower model complexity, where the optimal number of principal components is examined using sequential forward procedure. It also examines the minimum number of electrodes placed on the site which has greater impact on stress detection. They showed that using only 58 and 15 principal components, the accuracy of detecting stress reached 100% and 99.8% using cubic SVM and KNN classifiers respectively whereas our study showed an accuracy of **85**.**31%** and **83**.**44%** with SVM and KNN respectively.

But machine learning applied to the good and bad performers to identify a marker couldn’t yield a similar statistically significant biomarker. Possibly the sample size was small to achieve a significant discriminating biomarker. Another limitation of the study is that for each arithmetic calculation load the amount of change in the feature couldn’t be quantified because of the gross variation for each subject in the calculation they could perform.

## Conclusion

Machine learning by SVM revealed that ***Log(psd of alpha/psd of gamma)*** can differentiate mental arithmetic task and rest states with eyes closed. Frontal lobe is affected more widely and deeply during mental arithmetic task cognitive load. Entropy and fractal dimension decreased in the frontal lobe proportional to the cognitive load.

## Methods

### Participants

The dataset was obtained from the PhysioNet “EEG During Mental Arithmetic Tasks” database contributed by contributed by Igor Zyma, Sergii Tukaev, and Ivan Seleznov, National Technical University of Ukraine “Igor Sikorsky Kyiv Polytechnic Institute”, Department of Electronic Engineering^32,33^.(Zyma et al, Goldberger et al physiobank)

Totally, 66 healthy right-handed volunteers (47 women and 19 men) were initially involved in the study. All participants are 1st–3rd year students of the Taras Shevchenko National University of Kyiv (Educational and Scientific Centre “Institute of Biology and Medicine” and Faculty of Psychology) aged 18 to 26 years (Mean = 18.6 years, Standard Deviation (SD) = 0.87 years). The details of the participants are described in Table 1.

**Table 1.**
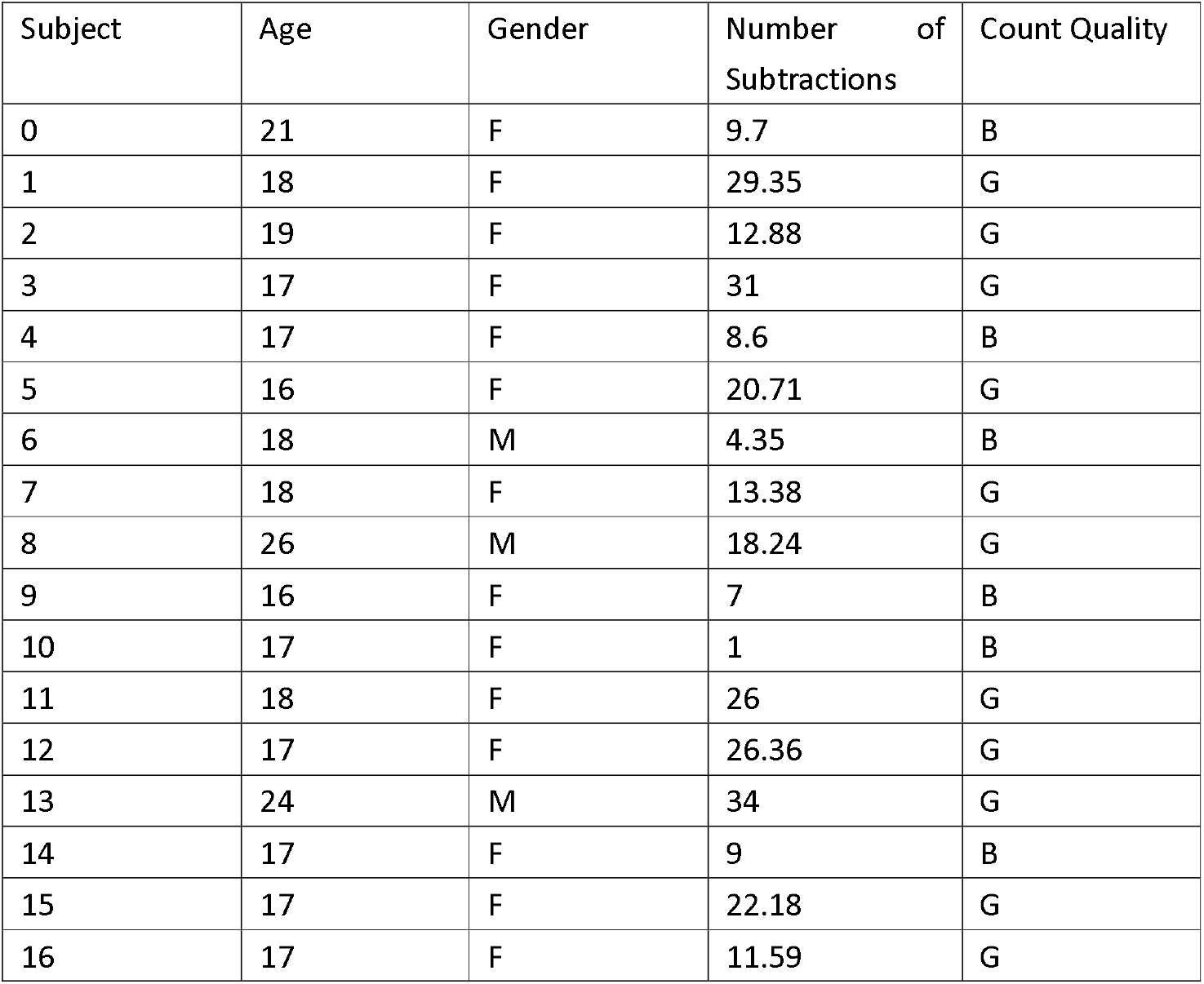
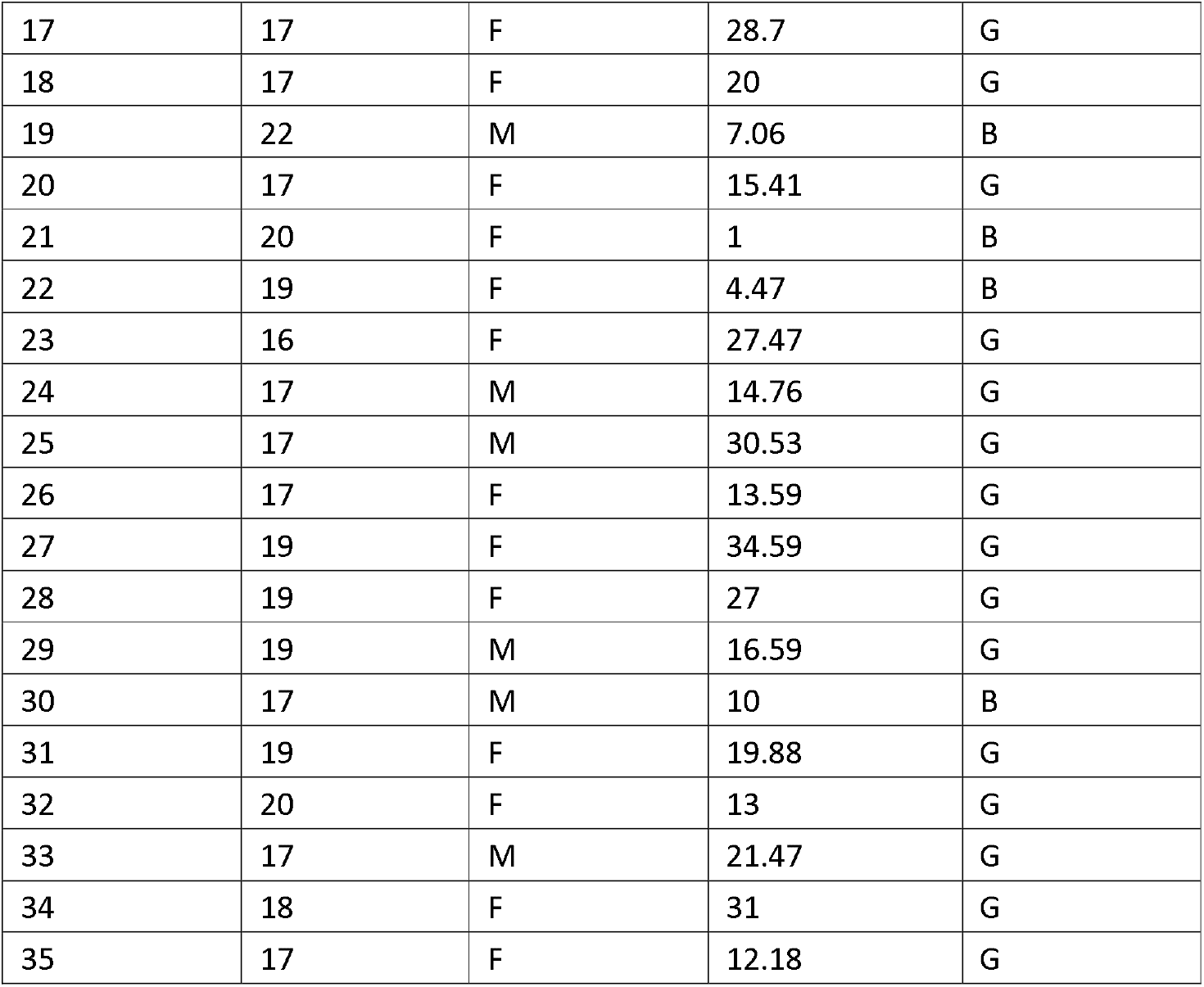
Participant details

The participants were eligible to enroll in the study if they had normal or corrected-to-normal visual acuity, normal color vision, and had no clinical manifestations of mental or cognitive impairment or verbal or non-verbal learning disabilities. Exclusion criteria were the use of psychoactive medication, psychiatric or neurological complaints and drug or alcohol addiction.

The study was approved by the Bioethics Commission of Educational and Scientific Centre “Institute of Biology and Medicine”, Taras Shevchenko National University of Kyiv (Conclusion from 15 August 2018, project title “Detrended fluctuation analysis of activation re-arrangement in EEG dynamics during cognitive workload”). Each subject signed written informed consent following the World Medical Association (WMA) declaration of Helsinki of 1975 (http://www.wma.net/Data 2019, 4, 14 5 of 6 en/30publications/10policies/b3/), revised in 2008, the Declaration of Principles on Tolerance (28th session of the General Conference of UNESCO, Paris, 16 November 1995), the Convention for the protection of Human Rights and Dignity of the Human Being with regard to the Application of Biology and Medicine: Convention on Human Rights and Biomedicine (Oviedo, 4 April 1997).

### Apparatus/materials

The entire analysis is summated in Figure 1. Neurocom monopolar EEG 23-channel system (Ukraine, XAI-MEDICA) was used to obtain the recording. Silver/silver chloride electrodes were placed on the scalp at symmetrical anterior frontal (Fp1, Fp2), frontal (F3, F4, Fz, F7, F8), central (C3, C4, Cz) parietal (P3, P4, Pz), occipital (O1, O2), and temporal (T3, T4, T5, T6) recording sites as per the **10-20 International** System of Electrode Placement. All electrodes were referenced to the interconnected ear reference electrodes. The inter-electrode impedance was below 5 kΩ and sample rate 500 Hz per channel.

**Figure 1.**
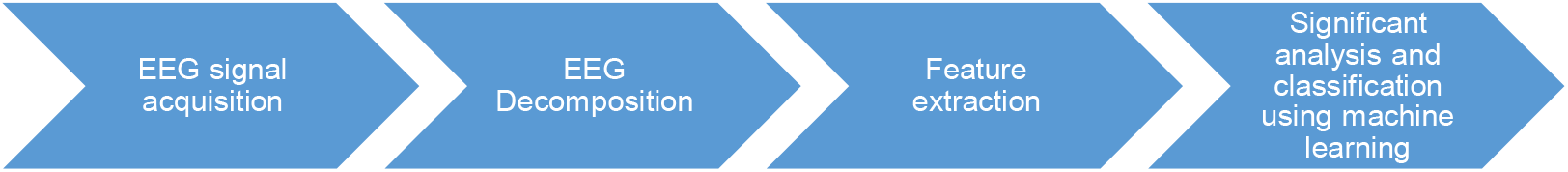
Workflow of the analysis.

Each recording consists of an artefact-free EEG segments of 180 s for resting state and 60 s for task state. 30 of the 66 initial participants were excluded from the database due to poor EEG quality by a board certified electro neurophysiologist, hence the final number of 36 subjects.

### Procedure

Tasks in this study involved the serial subtraction of two numbers. Each trial started with the oral communication of the 4-digit (minuend) and 2-digit (subtrahend) numbers (e.g., 7456 and 45, 2343 and 48, etc.). Serial subtraction during 15 min is considered to be a psychosocial stress^34^. In this way, our study design required intensive cognitive activity from the subjects. Intensive mental load is accompanied by a change in the emotional background when the subject makes additional effort to resolve tasks, so one can talk about evoked emotions in this case. During EEG recording, the participants sat in a dark soundproof chamber, comfortably reclined in an armchair. Serial subtraction is a composite measure of auditory attention/ concentration, mental tracking, and computation. Hence to avoid all distractors, before the experiment is begun, participants were instructed to try to relax during the rest state. Then they were asked to count mentally without speaking or using finger movements, accurately and quickly, in the rhythm they had determined. After 3 min of adaptation to experimental conditions, EEG registration of the rest state with closed eyes was made (over the next 3 min). Then the participants performed a mental arithmetic task—serial subtraction—for 4 min. Participants were interviewed about their strategies and experience after the experiment. These periods were selected since the task performance strategy is being formed simultaneously as the task is executed, and the emotional state of the participants is changing considerably due to intellectual overload.

### Data processing and analysis

EEG Pre-processing. EEG data were first exported to EEGLAB^35^ then analyzed mainly using MATLAB (R2018b). The length of rest state EEG signal is 180 seconds and the length of mental arithmetic EEG signal is 60 seconds. They were divided into 10-seconds epochs. However, the rest state EEG signal of subject 04 and subject 31 are shorter than other. Therefore, there are 851 epochs including 635 rest epochs and 216 task epochs.

High pass filter, low pass filter and band stop filter was applied in this dataset. 0.5Hz high pass filter was used to remove baseline. 45Hz low pass filter was used to remove high frequency noise. 50Hz notch pass filter to remove power frequency noise. The Independent Component Analysis(ICA) method was used to eliminate the artifacts (eyes, muscle and cardiac overlapping of the cardiac pulsation). The wave and power spectral density (PSD) of clean pre-processed data was shown in Figure 2. There are 19 channels, including FP1, FP2, F3, F4, F7, F8, T3, T4, C3, C4, T5, T6, P3, P4, O1, O2, Fz, Cz and Pz.

**Figure 2.**
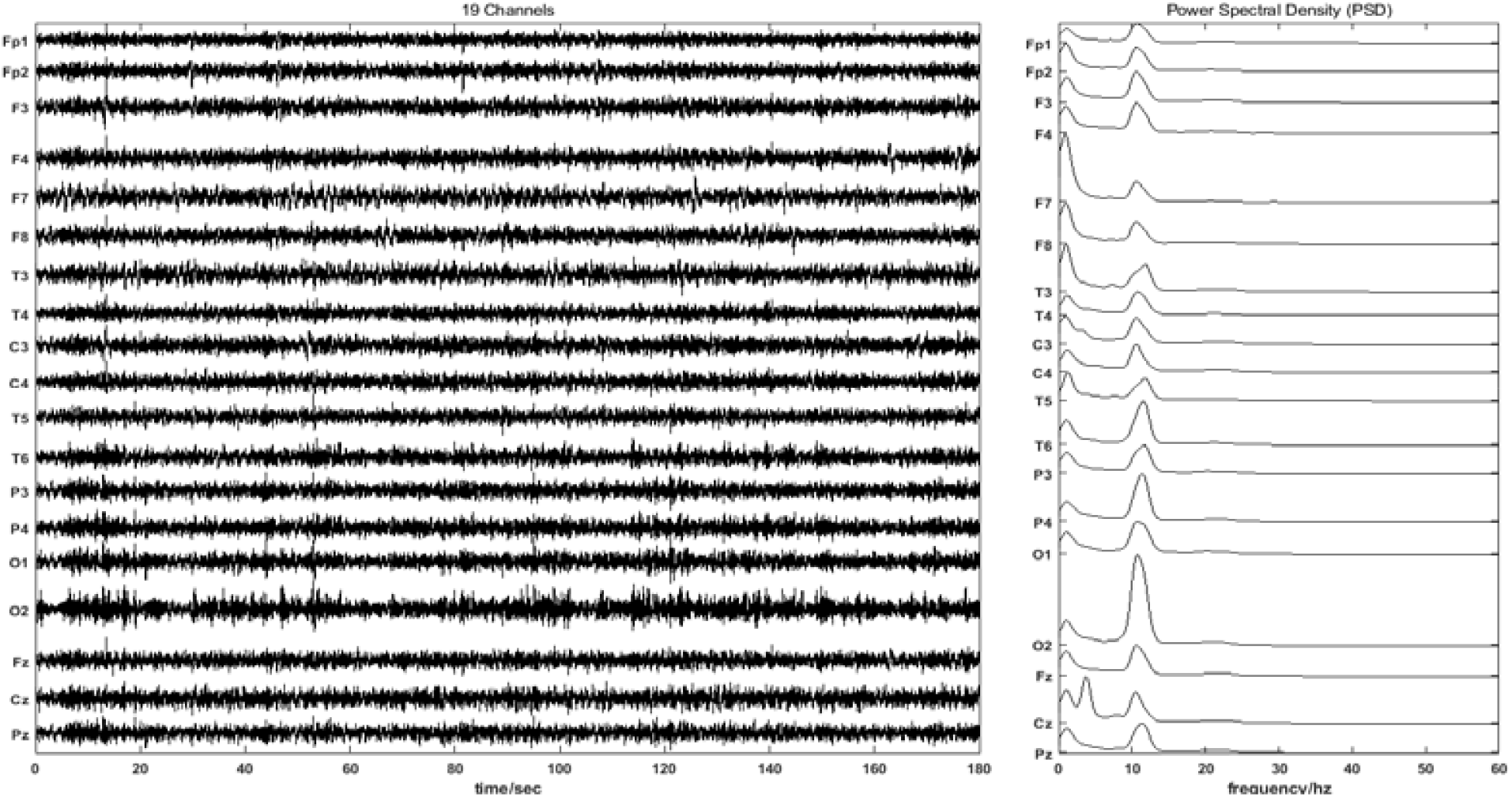
Processed signals of 19 channels with the estimated corresponding power spectral densities.

### Decomposition

The EEG data was decomposed into several frequency bands (delta, theta, alpha, beta and low gamma) based on wavelet packet transform with wavelet basis ‘sym7’ and level 7 using the Wavelet Toolbox™ software carried out in MATLAB (R2018b) (MathWorks Inc., Natick, MA). According to the following formula^36^(Daubechies), the decomposition was carried out and the results were obtained as shown in Table 2. and Figure 3.

**Table 2.** Decomposition of the EEG data: Frequency range of corresponding bands.

| Frequency Range | Name |
| --- | --- |
| 0.5000-3.9063Hz | delta |
| 5.8594-7.8125Hz | theta |
| 9.7656-13.6719Hz | alpha |
| 15.6250-29.2969Hz | beta |
| 31.2500-45.0000Hz | Low gamma |

**Figure 3.**
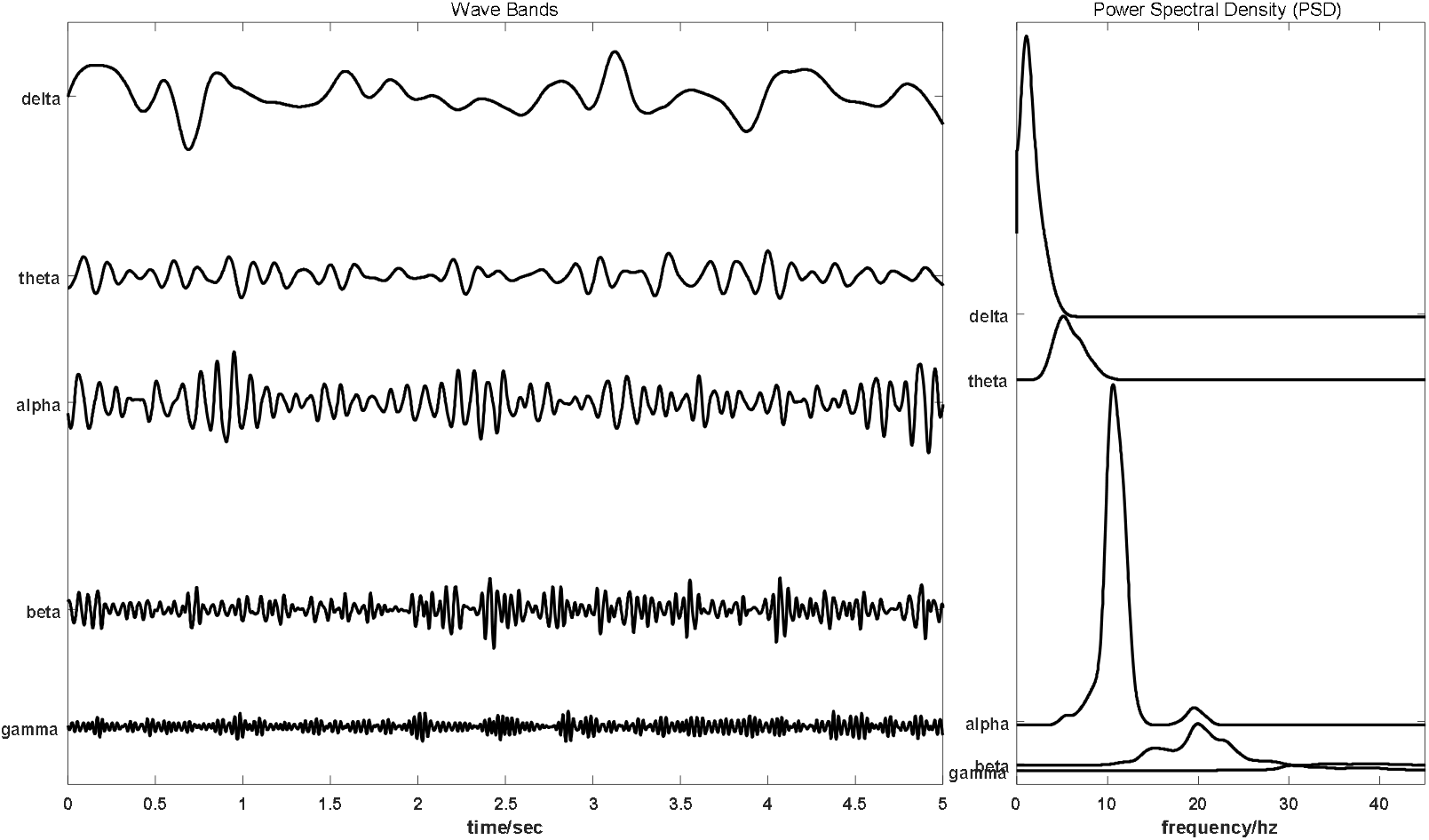
Decomposition of the EEG data into different frequency bands.

### Feature Extraction

In this study, the power spectral density (PSD), sample entropy (SE), fuzzy entropy (FE) and fractal dimension (FD) were used on five frequency bands (delta, theta, alpha, beta and low gamma) for rest and task states.

### Power spectral density

*Welch’s method*^37^ was used for estimating power spectra. It is carried out by dividing the time signal into successive blocks, forming the periodogram for each block, and averaging.

The **m** th windowed, zero-padded frame from the signal **x** is denoted by

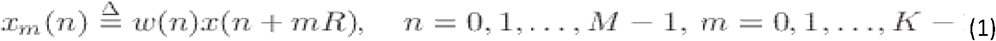

where *R* is defined as the window hop size, and let *K* denote the number of available frames. Then the periodogram of the **m** th block is given by

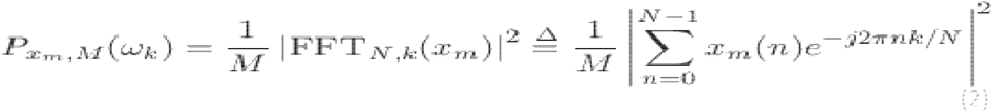

as before, and the Welch estimate of the power spectral density is given by

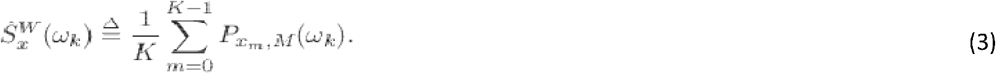

In other words, it’s just an average of periodograms across time. When **w(n)** is the rectangular window, the periodograms are formed from non-overlapping successive blocks of data.

### Sample entropy

Sample entropy is a modification of approximate entropy^38^. We have an imfN=xi=(x1,x2,x3,…,xN) i=1,2,3,…,N and use a time interval to reconstruct series Xm(i)=(xi,xi+1,xi+2,…,xi+m−1) i=1,2,3,…,N−m+1. The length of sequence is m. The distance function of two sequences is d[Xm(i),Xm(j)]. For a given embedding dimension m, tolerance r and number of data points N, SE is expressed as:

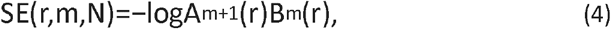

where, Bm(r) is the number of template vector pairs having d[Xm(i),Xm(j)]<r and represents the similarity between two sequences of length *m*, Am+1(r) is the number of template vector pairs having d[Xm+1(i),Xm+1(j)]<r and represents the similarity between two sequences of length *m* + 1.

### Fuzzy Entropy

Fuzzy entropy is the entropy of a fuzzy set, which loosely represents the information of uncertainty^39,40^

For an imfN=xi=(x1,x2,x3,…,xN) i=1,2,3,…,N, we reconstruct series with length *m*:

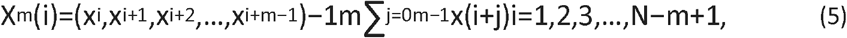

The distance function dmij of two sequences is d[Xm(I),Xm(j)]. Given *n* and *r*, calculate the similarity degree Dmij through a fuzzy function μ(dmij,n,r).

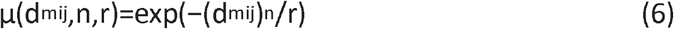

Define the function ∅m as

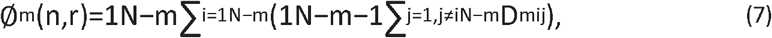

∅m+1(n,r) is got similarly.

Lastly, the FuzzyEn(m,n,r) of the series is shown in equation (8).

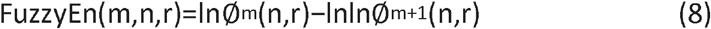

### Fractal dimension

Fractal dimension as one of the chaos measurement tools is widely utilized in the determination of chaotic behavior of signals. We have used the box-counting method for the FD calculation. In this method, a network of squares is created on the contour, and the number of squares in this network that includes a part of the curve is calculated. The size of the squares changes, then, the occupied squares are counted again. A point is obtained by calculating the logarithm of the number of counted squares and the logarithm for the reduction coefficient. New points are obtained by reducing the size of the network, which is connected in a curve. Calculating the slope of this curve indicates the FD. Ahmadlou et al compared two main algorithms for calculating fractal dimension from EEG. HFD was shown to provide better discrimination (91.3%) compared to Katz’s Fractal Dimension^41^. In this paper we have used the Higuchi’s FD(Higuchi) due to its high level of accuracy^42^.

Equation (9) describes how the self-similarity dimension (D), the number of self-similar pieces (a) and the reduction factor (1/S) are related together.

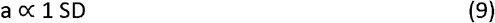

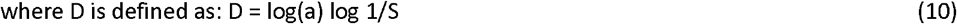

D can be calculated by estimating the slope of the approximated line for the plot of log (a) vs. log (1/s).

### Statistical software

All analyses were carried out in MATLAB (R2018b) (MathWorks Inc., Natick, MA). All reported statistical tests in the present study are two-sided tests wherever applicable. The significant difference was defined as the p-value < 0.05. Parametric test ANOVA1, which is a one-way analysis of variance, was used for the data that satisfied the assumptions. Otherwise, a nonparametric Kruskal-Wallis test was used.

## Data Availability

Te datasets analysed during the current study are available from the corresponding author on reasonable request.

